# Hand washing at critical times and associated factors among mothers/caregivers of under-five children in Nefas Silk Lafto Sub-City, Addis Ababa, Ethiopia, 2019: Facility based cross-sectional study

**DOI:** 10.1101/2022.06.24.22276847

**Authors:** Ermias Wabeto Wana, Nardos Anbese Mengesha

## Abstract

**Background:** Hand washing is the simplest, most affordable and effective means of limiting spread of infections. It has especial importance for children because they are more susceptible to infections acquired from unwashed hands. Despite increasing efforts to improve hand washing at critical times, mothers/caregivers of under-five children fail to practice; but, the reason is unclear. Thus, this study was aimed to identify the magnitude and factors associated with hand washing at critical times among mothers/caregivers of under-five children.

**Methods:** An institution based cross-sectional study was conducted in Nefas Silk Lafto Sub-City by April 2019, and 312 mothers/caregivers were participated. The data were collected by interviewing mothers, entered and analyzed with statistical package for social science 20 (SPSS-20). The findings were presented with crude odds ratios (COR) and Adjusted Odds Ratios (AOR) with their respective 95% confidence intervals (CI). A P-value less than 0.05 was used statistical significance level.

**Results:** 232 (74.4%; 95% CI [69.6%-79.2%]) mothers/caregivers washed their hands at critical times. Illiterate mothers had 66% reduced (AOR= 0.34; 95%CI [0.17-0.69]) odds of washing hands at critical times than literate mothers. Mothers who did not own tap water in their back yard had 62% (AOR= 0.38; 95%CI [0.18-0.80]) reduced odds of hand washing at critical times than their counterparts. As compared to the mothers from the poorest households, those from middle, richer and the richest households had 4.56 (AOR= 4.56; 95%CI [1.84-11.33]), 5.61 (AOR= 5.61; 95%CI [2.11-15.30]) and 6.14 (AOR= 6.14; 95%CI [2.24-16.72]) times increased likelihood of washing hands at critical times.

**Conclusion:** Three fourth of mothers practiced hand washing at critical times, and improving maternal literacy, household economy and availability of water source in backyard are needed to maintain and enhance the practice.

## Background

Hand washing is an easy and do yourself and affordable task for every community. It is an effective means of stopping or limiting the spread of infections via feces, body fluids, and inanimate objects (1). Hand washing with a soap and water is one of the most effective measures against infectious diseases like diarrhea (2, 3). It is an easy to do, safe, cheap and not time consuming, and it can be effectively adopted by any socio-economic class and any community (4, 5). It’s also culture-sensitive and generally acceptable across many population groups (3). Consistent practice of proper hand washing with soap and water has great potential of keeping a family, especially mothers and children away from germs and hence ill health. Mothers/caregivers engage in different activities like cleaning child’s bottom, clean the home compound and environment, have contact with domestic animals and visit toilet that interacts hands with micro-organism; thus, they are expected to properly wash their hands at critical moments (4).

Globally, about 1.7 billion cases and 525,000 deaths of under-five children occur per year due to diarrhea secondary to poor hygiene (6). In developing countries, 80% of the diseases burden is associated with poor domestic and personal hygiene (7). More than 2 million people, mostly children die yearly due to diarrhea; the same numbers of children also die from acute respiratory infections (8). In Ethiopia, more than 250,000 children death per year and 60% to 80% of all deaths were attributed to poor sanitation and hygiene, and 70,000 under-five deaths per year in 2018 were due to diarrhea only (9-11).

Five moments are considered as critical times to wash hands; after defecation, after handling child/adult feces or cleaning child’s bottom, after cleaning the environment, before preparing food and before eating food (4, 12-14).

Evidences also show that hand washing at critical times reduces rates of diarrheal diseases like cholera and dysentery by 41%(15). It also has better reward; for example 3.35 dollar investment in hand washing gives backward benefit of 11.0 dollar investment in latrine construction, 200 dollar investment in water supply and millions of dollars investment in immunization; but, proper hand washing with water and soap at critical times is remained at lower proportion, zero to 34% (16). According to the World Health Organization (WHO) and the United Nations Children’s Fund (UNICEF) joint monitoring program estimate on household drinking water, sanitation and hygiene, only 8% of Ethiopians had basic hand washing practice (with water and soap or substitutes) and the richest and urban residents practiced better hand washing than their counterparts (17).

A study conducted in debark town, Amhara regional state in 2018 showed that 52.2% of study participants practiced hand washing with water and soap/substitutes at critical times and the practice was enhanced by desirable attitude, presence of water for washing hands and a good knowledge of hand washing and its benefits (18).

A study conducted in Wondogenet woreda, Oromia region showed that 87% of mothers of under five children practiced hand washing at critical times and their practice was enhanced by maternal literacy (19). Another study carried out in Bechi-Maji Zone, in SNNPR showed that 34.6% of the participants practiced hand washing at critical times and literate mothers had 90% reduced odds of practicing hand washing at critical times (20).

Studies in Ethiopia used insignificant moments (after food preparation) or practically inconvenient moments like washing hands after sneezing, coughing and touching clothes as critical times and others used four or five moments and revealed inconsistent findings on level and factors associated with hand washing at critical times among mothers of under-five children. Besides to this, a little is known about the practice among mothers of under-five children in Nefas Silk Lafto Sub-City (NSLSC), Addis Ababa, Ethiopia. Therefore, the current study aimed at assessing hand washing practice at critical times (after visiting toilet, after cleaning child’s bottom, before preparing food, before eating/feeding and cleaning compound) and associated factors among mothers of under-children in NSLSC, Addis Ababa, Ethiopia.

## Methods

### Study Area

NSLSC is one of the 10 Sub Cities in Addis Ababa city, the capital of Ethiopia. It borders with the Kolfe Keranio, Lideta, Kirkos, Bole and Akaki-Kaliti sub-cities. The Sub City is located between coordinates 8°56’57” latitude and 38°43’58” longitude. It is divided into thirteen woredas and has total area coverage of 68.30 km2 (26.37 square miles) hectares of land. The total population of the sub-city was estimated to be 366,006 with male to female ratio of 1to1 in 2021. A population is being served by ten health centers and two public hospitals, thirty four private clinics and health coverage of the sub-city is 100% (21). Regarding the water supply, about 95% of household get water from an improved source (piped water sources) and estimated domestic per capita water consumption was 52 l/c/day. An improved sanitation coverage was below ten percent (8%) in the sub-city and a hygiene index was medium (53%) (22), and there were no sustainably functioning public hand washing facilities in the sub-city.

### Study design and period

Institution based Cross sectional study was carried out in December, 2019.

### Source and study populations

A Source population is all mothers/caregivers of under-five children living in NSLSC; while study populations were who visited health facility during the survey period.

### Sample size determination and sampling technique

The sample size was calculated manually using the single population proportion formula with the following assumptions; the expected proportion of hand washing practice at critical times 89.6% (19), 95% significance level, 5% margin of error, 2 design effect and 10% expected non response rate gave us sample size of 315.

### Sampling procedures

NSLSC was selected from ten sub-cities in the city (Addis Ababa) due to its convenience to investigators. Four woredas (woreda one, two, four and twelve) were randomly selected by lottery method. The total sample required from each woreda was allocated proportionally to the size of mothers/caregivers of the under-five children in each woreda. Data were collected from mothers/caregivers visiting one randomly selected health center in each woreda (one, two, four and twelve). Mothers/caregivers visiting immunization clinics and visiting health center for seeking medical service for their children were selected at interval of three via systematic random sampling technique.

### Study variables

hand washing at critical time was the dependent variable whereas maternal literacy, paternal literacy, availability of tap water in the back yard, access to health education by mothers/caregivers on hand washing, maternal knowledge on purpose of hand washing at critical times, average family size, number of living children the mother gave birth to and household wealth status were explanatory variables.

### Data collection

Data were collected by pre-oriented certified nurses with diploma in academic qualification with a strong supervision and follow up by supervisor and the investigators. A pretested structured questionnaire was used to collect the data and interview was administered to the respondents in an Amharic language. One day orientation was given to the data collectors and a supervisor on objective of the study, how to approach and interview the participants and keep the quality of the data. Before collecting the actual data, 5% of the sample size was pretested on other health facilities to validate the tool and the necessary correction was made on data collection material. Data on socio-demographic/economic, information/knowledge related to hand washing and hand washing practice of mother/caregiver were collected on a daily basis during the month December 2019.

### Operational definitions

Hand washing at critical times: women had proper hand washing at critical times if she washed hand with water and soap or substitutes after using the toilet, cleaning a child’s bottom, before preparing and eating food and after cleaning compounds [(4, 13, 14)].

Mother/caregiver: we considered biological mothers and others responsible for caring a child whose biological mother was died or can no longer care for a child due to any reason as mother/caregiver.

Under-five Children: children birth to 59 months old were called as under-five children Wealth Status of the household: wealth status created by Principal component analysis and household belonged to first, second, third, fourth and fifth quintile were categorized as poorest, poor, middle, rich and the richest, respectively.

Maternal and paternal literacy: Mothers or fathers who have achieved education from simply reading and writing to higher degree were considered as literate; while those who at least cannot read and write were considered as illiterate.

### Data analysis

The educational status of mothers/caregivers was categorized into illiterate and literate because some of the expected cells violated chi-square assumption (contained value less than five) when the highest grade point achieved was assumed. The wealth index (indicator of living standard of house hold) was constructed through principal component analysis (PCA) from household assets. We ranked the extracted component into quintiles (five tiles); each quintile holding 20% of households, and the household that belongs to the first quintile was categorized as the poorest and the households that belong to the second, third, fourth and fifth quintiles were categorized as poor, middle, rich and the richest, respectively. The value/score one was assigned for a moment when a mother/caregiver had washed her hands with water and soap/substitutes and zero was assigned if a mother/caregiver did not. The scores were summed up to 5 and a mother/caregiver who had a total score of 5 was considered as washed her hands at critical times. If the sum of scores is less than five, a mother/caregiver was treated as did not wash her hands at critical moments. Descriptive statistics had been used to describe the study variables and logistic regression was undertaken to identify the factors associated with hand washing practices.

A multi-collinearity test was carried out to check interrelationship between an independent variables with variable inflation factor less than ten and a chi-square test was conducted to check the expected cells adequacy. Findings of descriptive analysis were presented by frequency and percentage, mean or median and the regression out puts were presented by crude and adjusted odds ratios with their 95% confidence intervals. A p-value of less than 0.05 in multivariable analyses was taken as the cut-off point for statistical significance.

### Quality assurance

Before a collection of the actual data, 5% of total sample were pretested and necessary corrections were made accordingly. Translation and back translation of questionnaire from English to Amharic and back to English by different individuals to check consistency was undertaken. At the end of each data collection day, data were checked for completeness and consistency, and discussion with the research assistants was carried out. Unique identification number was assigned to each questionnaire after editing, checking completeness and consistency. After entering the data into SPSS 20, 10% sample was checked for correct entry and cleaning was also continued up to the end of descriptive analysis.

## Results

Three hundred and twelve mothers/care givers of children were participated in the study and this gave the response rate of 99%. The median age of the respondents was 28 (IQR=4) years and minimum and maximum ages were 18 and 50 years in that order. Regarding educational attainment of respondents, 94 (30.1%), 137 (43.9%) and 12(3.9%) attained primary (grade 1-8); high school and certificate and above, respectively (Table1).

**Table 1:**
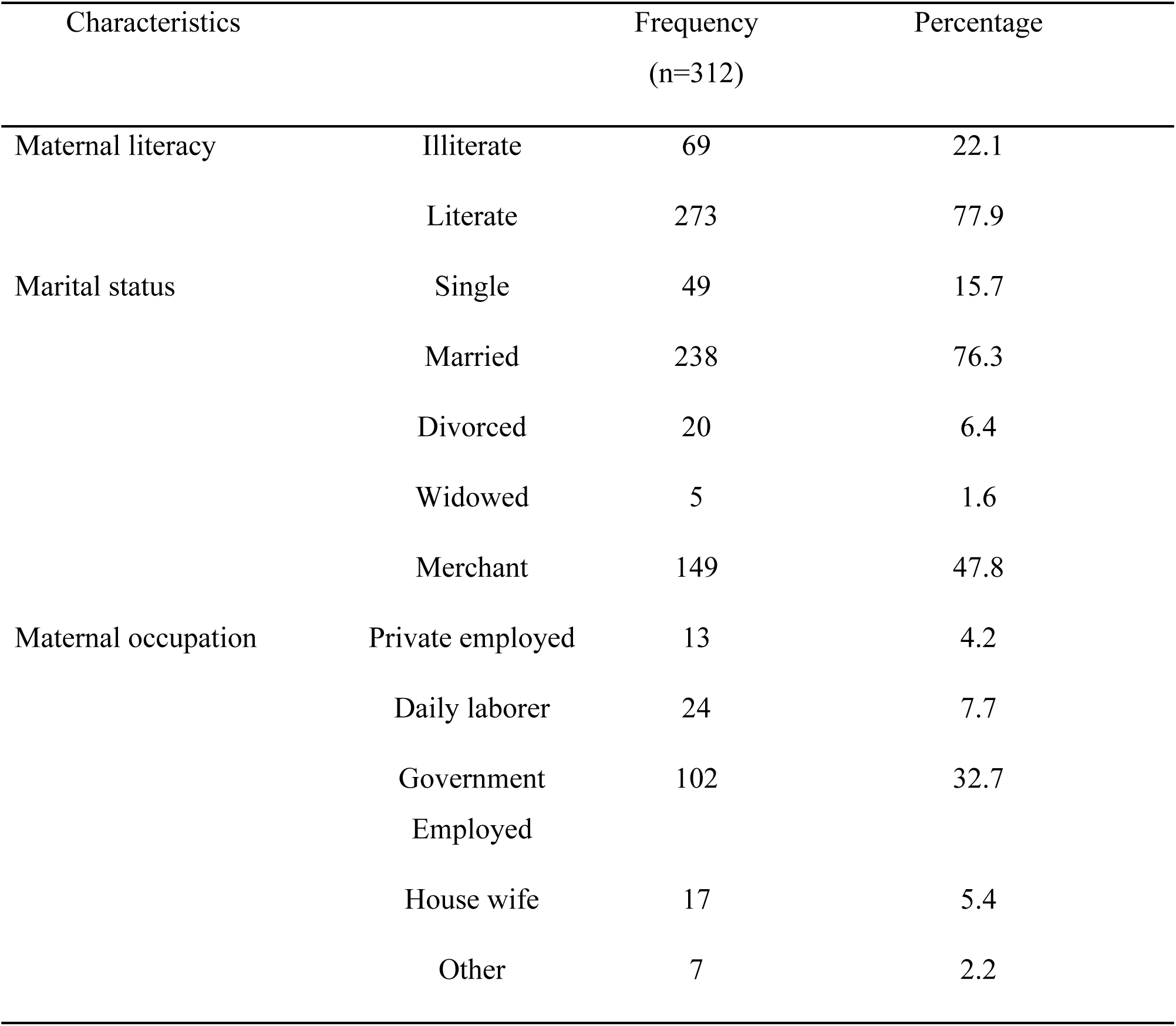
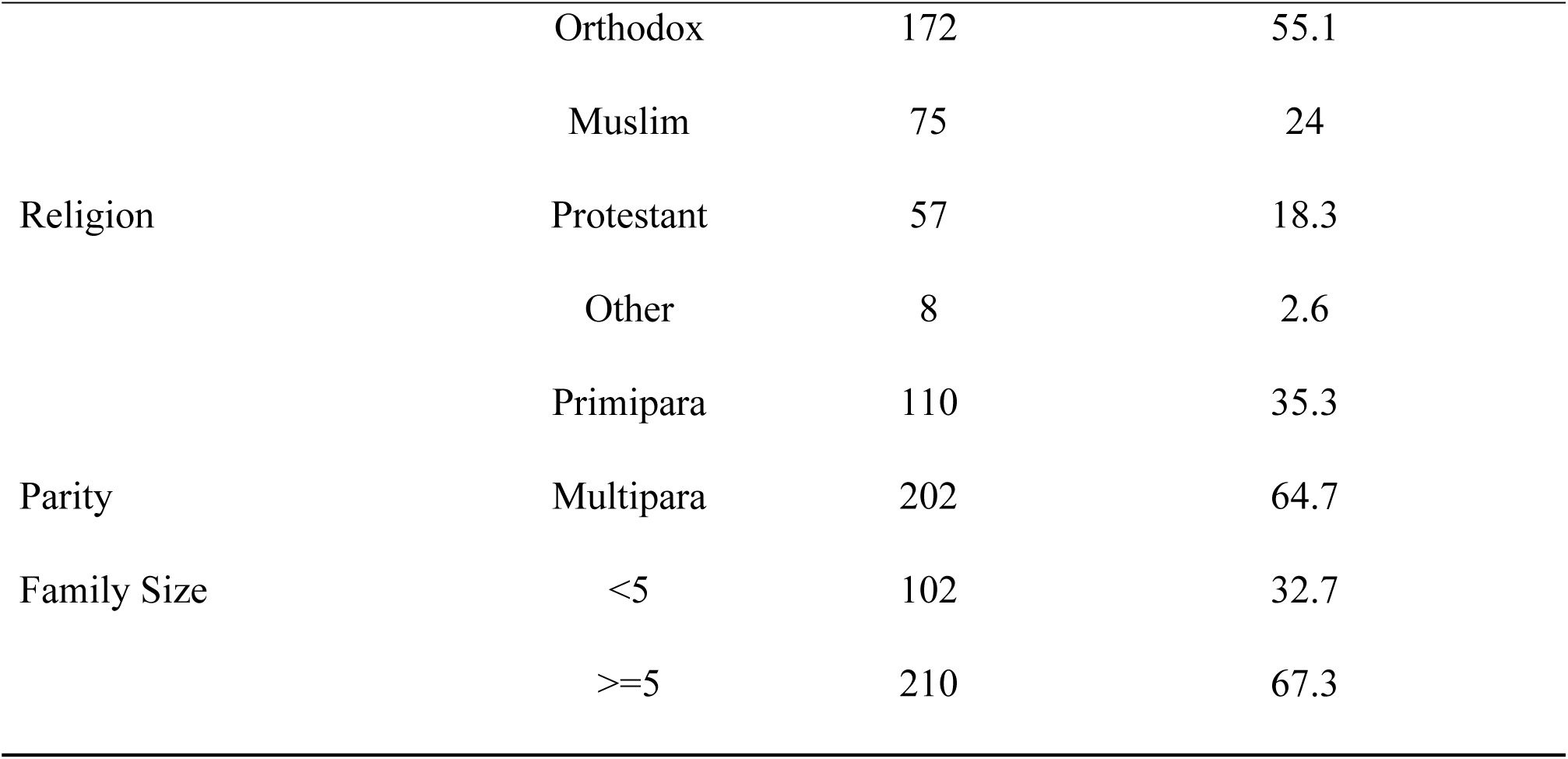
Socio-demographic characteristics of respondents in NSLSC, 2019

| Characteristics |  | Frequency<br>(n=312) | Percentage |
| --- | --- | --- | --- |
| Maternal literacy | Illiterate | 69 | 22.1 |
|  | Literate | 273 | 77.9 |
| Marital status | Single | 49 | 15.7 |
|  | Married | 238 | 76.3 |
|  | Divorced | 20 | 6.4 |
|  | Widowed | 5 | 1.6 |
|  | Merchant | 149 | 47.8 |
| Maternal occupation | Private employed | 13 | 4.2 |
|  | Daily laborer | 24 | 7.7 |
|  | Government<br>Employed | 102 | 32.7 |
|  | House wife | 17 | 5.4 |
|  | Other | 7 | 2.2 |

Table 1 continued
|  |  |  |  |
| --- | --- | --- | --- |
|  | Orthodox | 172 | 55.1 |
|  | Muslim | 75 | 24 |
| Religion | Protestant | 57 | 18.3 |
|  | Other | 8 | 2.6 |
|  | Primipara | 110 | 35.3 |
| Parity | Multipara | 202 | 64.7 |
| Family Size | <5 | 102 | 32.7 |
|  | >=5 | 210 | 67.3 |

Concerning the Economic status as indicated by house hold wealth index, 63 (20.2%) fall under poorest category, 90(28.8%) poor, 59(18.9%) middle, 47 (15.1%) rich and 53 (17.0%) fall in the richest economic group. More than three fourth (77.9%) of the fathers were at least able to write and read.

### Information and knowledge regarding Hand washing

More than three fourth (84.3%) of the respondents had ever heard of hand washing and 164 (52.6%) of them got information on hand washing at the critical times in the last one year. Information sources were health workers; 150 (56.9%), schools; 47 (17.9%), TV/Radio; 28 (10.5%), religious leaders or church; 22 (8.2%), neighbors/friends; 6(2.2%) and other sources; 10 (3.2%). More than half of respondents, 215 (69%) were washing hands to prevent illness and or infestation by germs. Thirty four (10.8%) of the study participants thought that hand washing is trustworthy to prevent disease for the entire community and 14(4.5%) mentioned that washing hands gives sense of cleanliness and confidence. Respondents pointed out, disease occurrences can be prevented by hand washing; 92 (29.5%), 195(51.0%) and six (1.9%) of them mentioned that typhoid, diarrhea and respiratory problem, respectively can be prevented by proper hand washing.

### Hand washing practice

Three hundred and twelve of the respondents (100%) reported that they used to wash their hands as needed (Table 2).

**Table 2:**
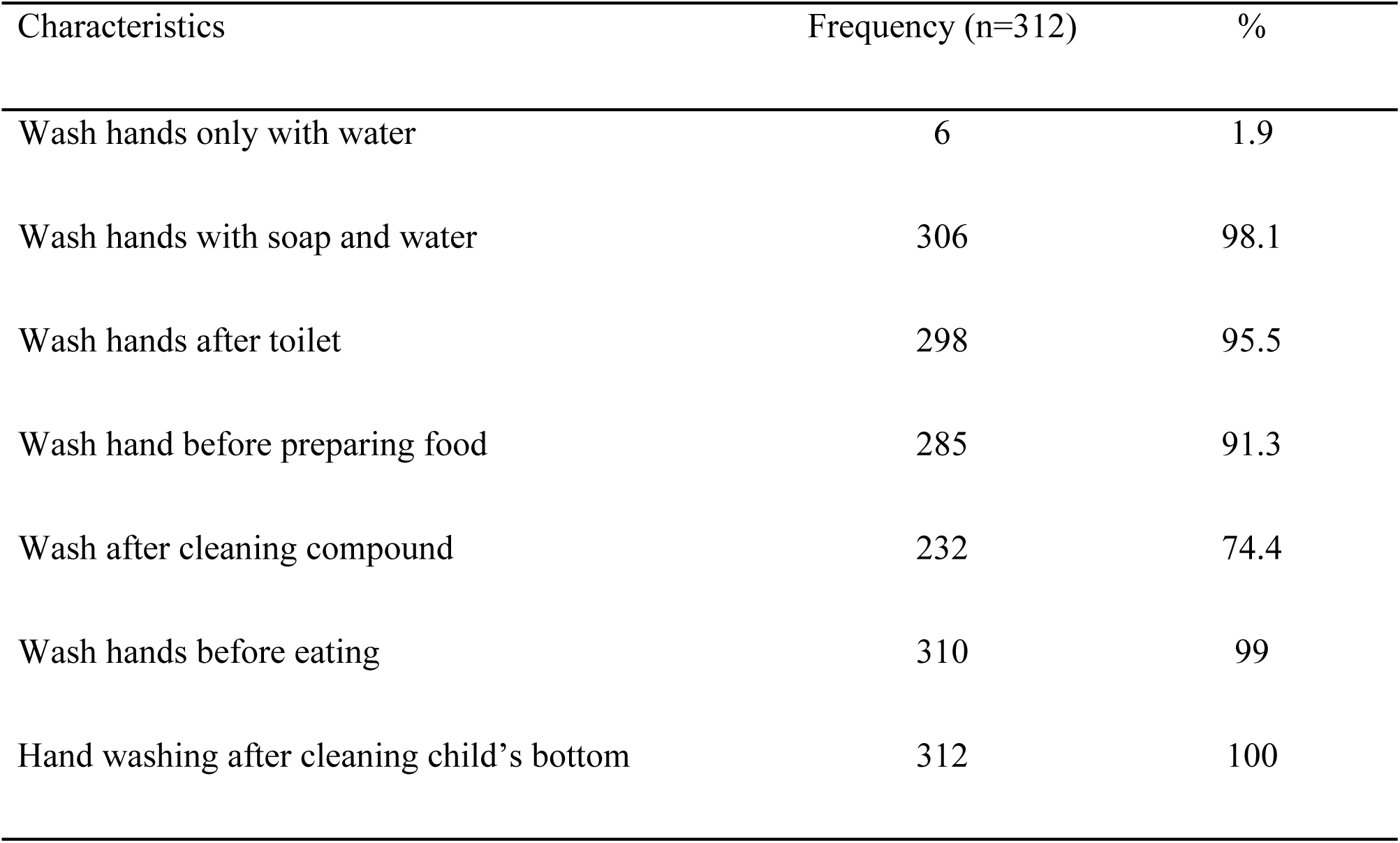
Hand washing practices of mothers of children in NSLSC, Addis Ababa, 2019

Among mothers of children who participated in the study, 232 (74.4%; 95% CI [69.6%-79.2%]) practiced hand washing at critical times during study period. Regarding water sources for household domestic purposes, majority of participants, 278 (89.1%) were using tap water for their domestic purpose. The remaining 14 (4.5%), 11 (3.5%) and nine (2.9%) of households mentioned that they were using rain water, spring and dug well, respectively. Mothers/caregivers in the study area reported that they had faced problems related with hand washing practice at critical times with water and soap; 114 (36.5%) faced inadequate water supply, 165 (52.9%) could not find soap when needed, 27 (8.7%) had inadequate time to wash hands at critical times and six (1.9%) had faced other challenges.

### Factors associated with washing hands at critical times

Eight variables (maternal literacy, paternal literacy, availability of water in the back yard, access to health education by mothers/caregivers on hand washing, maternal knowledge on purpose of hand washing at critical times, average family size, number of living children the mother gave birth to and household wealth status) were included in bi-variable binary logistic regression and six variables (maternal literacy, paternal literacy, availability of water source in the back yard, access to health education by mothers/caregivers on hand washing at critical times, maternal knowledge on purpose of hand washing at critical times and household wealth status) had p-value below 0.25 and became candidate for multivariable binary logistic regression model. Ultimately, three variables (maternal literacy status, availability of tap in the backyard of the household and household’s economic status as measured by wealth index) were found to be statistically significant determinants of hand washing at critical times among mothers/caregivers of under-five children in the study area. Illiterate mothers/caregivers had 66% reduced (AOR= 0.34; 95%CI [0.17-0.69]) odds of washing hands at critical times than literate mothers. Mothers/caregivers who did not own water supply in their back yard had 62% (AOR= 0.38; 95%CI [0.18-0.80]) reduced odds of hand washing at critical times than their counterparts. As compared to the mothers from the poorest households, those from middle, richer and the richest households had 4.56 (AOR= 4.56; 95%CI [1.84-11.33]), 5.61 (AOR= 5.61; 95%CI [2.11-15.30]) and 6.14 (AOR= 6.14; 95%CI [2.24-16.72]) times increased likelihood of washing hands at critical times (Table 3).

**Table 3:** Factors associated with hand washing at critical times among mothers in NSLSC, Addis Ababa, Ethiopia, 2019

| Variable |  | Wash hands at critical |  | COR with 95% CI | AOR with 95% CI |
| --- | --- | --- | --- | --- | --- |
|  |  | Times |  |  |  |
|  |  | Yes =232 | No =80 |  |  |
| Maternal literacy | Illiterate | 43 | 26 | 0.47(0.26-0.84) | 0.34(0.17-0.69)** |
|  | Literate | 189 | 54 | 1 | 1 |
| Paternal Literacy | Illiterate | 48 | 18 | 0.89(0.48-1.66) | 1.07(0.51-2.27) |
|  | Literate | 184 | 62 | 1 | 1 |
| Availability of tap water<br>in the back yard | No | 39 | 21 | 0.56(0.31-1.04) | 0.38(0.18-0.80)** |
|  | Yes | 193 | 59 | 1 | 1 |
| Mother Knows a benefit<br>of hand washing at critical<br>times | No | 18 | 3 | 2.16(0.62-7.53) | 2.00(0.50-7.89) |
|  | Yes | 214 | 77 | 1 | 1 |
| Mother Got Health<br>education on hand<br>washing during health<br>facility visit | No | 106 | 40 | 0.84(0.51-1.40) | 0.73(0.41-1.32) |
|  | Yes | 126 | 40 | 1 | 1 |
| Average Family size | ≥5 | 73 | 29 | 0.81(0.47-1.37) | 0.78(0.19-3.14) |
|  | <5 | 159 | 51 | 1 | 1 |
| Parity | Primi-<br>para | 80 | 24 | 1.20(0.67-2.03) | 1.05(0.65-2.66) |
|  | Multi-Para | 152 | 56 | 1 | 1 |

Table 3 continued
|  |  |  |  |  |  |
| --- | --- | --- | --- | --- | --- |
|  | Poorest | 36 | 27 | 1 | 1 |
|  | Poor | 62 | 29 | 2.16(1.14-4.10) | 1.27(0.63-2.60) |
| Wealth index | Middle | 50 | 9 | 5.70(2.45-13.29) | 4.56(1.84-11.33)** |
|  | Rich | 39 | 8 | 20.00(4.50-89.21) | 5.61(2.1-15.30)** |
|  | Richest | 45 | 7 | 15.79(4.49-55.29) | 6.14(2.24-16.72)** |

\*\* *Variables statistically significantly associated with hand washing at critical times*

## Discussion

The current study carried out on magnitude and factors associated with hand washing practice at critical times among mothers/caregivers of under-five children in NSLSC, Addis Ababa, Ethiopia. The study witnessed that about three fourth (74.4%) of respondents were practicing hand washing at critical times and their practice was affected by maternal literacy, availability of taped water and household economic status as indicated by a wealth index.

Out of three hundred and twelve participants involved in the study, 232 (74.4%) were washing their hands at critical times. The finding is lower than what was shown by a study conducted in Wondongent Woreda in Oromia Region which showed 87% (19). This may be due to less knowledge of respondents on diarrhea can be prevented by hand washing in a current case (51%) than 100% in Wondogent Woreda. Other possibility might be small area coverage (single kebele) by study carried out in Wondogenet woreda than the current case; where several kebeles or woredas participated and lower prevalence in some kebeles or woredas might have masked higher prevalence of hand washing at critical times among other kebeles/woredas in a current study area and this might have given a lower proportion of hand washing at critical times.

The finding shown by the current study also higher than finding witnessed by a study conducted in Benchmaji in May to August 2017 (20). The setting difference might affect the practice where the level of health intervention like health education and promotion and health related information and services dissemination are more easily accessed in the current setting than Benchmaji. Other reason might be the time interval between two studies during which attention is being given to hygiene as hygiene and sanitation related health problems account top position.

The current finding is higher than the finding (52.2%) shown by study conducted in Debark town, Amhara Regional state, Ethiopia (18) and the difference may be due to a study in Debark town used several moments (some such as after sneezing, touching clothes are inconvenient for hand) in addition five moments used in the current study. Other possibilities may be better access to basic services like water, soap and information regarding washing hands in the current setting than Debark town.

The current finding is higher than the finding showed by a study carried out in Nigeria in 2015; 30%(23). This difference may be justified by difference in proportion of household that own water supply (source) in their home; 53.9% of households own water supply in their home while majority of households in a current study (89.1%) had water source in their home or backyard. Other possibilities may be the time interval between the studies when different health interventions concerning hygiene and sanitation had been carried out and the setting difference; rural and urban residents were participated in the study in Nigeria, but, participants in the current study were solely urban residents. The finding is also higher than another study carried out in Nigeria in 2016 that revealed that only 32% of caregivers were observed to have good hand washing (24). The difference may be due to the presence of one extra parameter (washing hands before administering a drug to the child) and self-reporting in our case might result in exaggerated value while structured observations in the latter study might reveal a real practice.

The current finding showed a higher proportion of hand washing at critical times as compared to finding (40%) revealed by a study carried out in Pakistan (25). The inconsistency may be due to the difference in residence of respondents, 37.5% respondents were living rural in the latter study. The other possibility may be difference in a literacy rate among study participants (relatively a higher literacy rate in our case; 78% to 70%. The difference in access to protected water supply may also be another possibility; in a current study about 90% of the respondents were accessing water from tap while 74.5% of respondents in Pakistan had access to protected water source in general.

The current study witnessed that maternal literacy enhances hand washing practice at critical times. This is consistent with a studies conducted in Kirkos sub-city, Addis Ababa (26), Gondar comprehensive specialized hospital, Gondar town, North West Ethiopia (27), Wondogenet Woreda in 2014 (19) and WHO and UNICEF joint estimate on progress of household drinking water, hygiene and sanitation (17). But, the finding contrasts a study carried out in Benchmaji Zone Mizan town in 2017 where literate mothers had 90% reduced odds than their counterparts (20). The difference may be due to difference in the study settings; the current study was done in place where health related information can be easily disseminated to and accessed by the residents, and the current setting has a better access to water and other supplies required for hand washing.

The current study also showed that hand washing practice among mothers of the children is positively associated with the household’s economic status. The wealthier the household, the better hand washing practices at critical times and this agrees with WHO and UNICEF joint monitoring program estimate for 2000-2020 on progress of household drinking water, sanitation and hygiene that stated that wealthier and those live in urban setting had better basic hygiene than their counter parts (17). This finding also agrees with evidence showed by a study carried out in Holy family hospital, Rawalpindi, Pakistan (25). Wealthier households can afford purchasing of soap and can avail water at households even by buying.

Our study showed that availability of taped water at home or in backyard was directly related to hand washing at critical times and agrees with findings from Debark town which reported that availability of water affects good hand washing at critical times(18). This finding is also consistent with the information revealed by a study carried out in Nigeria (23) and (18).

## Study strength and limitations

### Strength

We have deployed the experienced enumerators to collect valid data, and close supervision was carried out by supervisor and investigators.

### Limitation

Since the proportion of hand washing at critical times reported under this study was self-reported, mothers/caregivers might have over reported their practice, and social desirability bias might have been introduced; thus, needs attention. The wider confidence intervals may be due to smaller sample size and indicates less stable power of the model, thus, precaution required. A cross-sectional study design captures snapshot and cannot show seasonal variation and temporal relationship; therefore, attention should be paid during generalization of the findings.

## Conclusion and Recommendations

About three quarter of mothers/caregivers practiced hand washing at critical times in the study area. The practice among mothers/caregivers was determined by maternal literacy, availability of tap water in the backyard or at home and economic status of the household. Thus, to maintain and enhance hand washing at critical times among mothers, the following actions should be implemented by respective bodies.

- NSLSC educational office should work hard to increase maternal literacy and educational attainment of mothers and girls for future.
- The water office of NSLSC is expected to improve access to tap to households
- The government as whole, NSLSC and other responsible bodies need to strive to improve economic status (wealth status) of economically disadvantaged households through income generating activities and social supports.
- We call on researchers to carry out similar study with larger sample size.

## Data Availability

The data set used during analysis of the current study is available with corresponding author and we ensure that any body can access it for scientific purposes

## Abbreviations

AURTI: Acute upper respiratory tract infection
MCH: Maternal and child health
NSLSC: Nefas Silk Lafto Sub-City
UNICEF: United Nation Children’s Fund
WASH: Water, sanitation and hygiene
WHO: World Health Organization

## Acknowledgements

We are very grateful towards Addis Ababa medical and Business College for allowing us to conduct this research work. Again our gratitude goes to study participants, data collectors and supervisors.

## Authors’ contribution

**EWW:** conceived and designed the study, supervised data collection, managed and analyzed the data, interpreted the findings and approved the final manuscript.

**NAM**: conceived and designed the study, supervised data collection, managed and analyzed the data, interpreted the findings and approved the final manuscript.

## Availability of data and materials

Data set used for analysis of the current study can be accessed from the corresponding author.

## Ethical Issues

Ethical approval was obtained from an institutional review board (IRB) of Addis Ababa medical and business college with reference number IRB/022/2011 according to the standardized principles and procedures in line with national and international guidelines. Written permission letter was received from NSLSC and respective randomly selected woredas. Oral consent was obtained from all the respondents after explaining the purpose of the study, risk/discomfort, and confidentiality of response, right to refuse and terminate participation in the study at any time. Address of contact person was disclosed for the participants to use when they have any doubt. Data was collected using codes and no name and personal private identifier had been used and the confidentiality of collected data was maintained.

## Consent for publication

Not applicable (No identifying images and personal details in the manuscript).

